# Protection of β cells against pro-inflammatory cytokine stress by the GDF15-ERBB2 signaling

**DOI:** 10.1101/2023.11.27.23298904

**Authors:** Soumyadeep Sarkar, Farooq Syed, Bobbie-Jo Webb-Robertson, John T. Melchior, Garrick Chang, Marina Gritsenko, Yi-Ting Wang, Chia-Feng Tsai, Jing Liu, Xiaoyan Yi, Yi Cui, Decio L. Eizirik, Thomas O. Metz, Marian Rewers, Carmella Evans-Molina, Raghavendra G. Mirmira, Ernesto S. Nakayasu

## Abstract

**Aim/hypothesis:** Growth/differentiation factor 15 (GDF15) is a therapeutic target for a variety of metabolic diseases, including type 1 diabetes (T1D). However, the nausea caused by GDF15 is a challenging point for therapeutic development. In addition, it is unknown why the endogenous GDF15 fails to protect from T1D development. Here, we investigate the GDF15 signaling in pancreatic islets towards opening possibilities for therapeutic targeting in β cells and to understand why this protection fails to occur naturally.

**Methods:** GDF15 signaling in islets was determined by proximity-ligation assay, untargeted proteomics, pathway analysis, and treatment of cells with specific inhibitors. To determine if GDF15 levels would increase prior to disease onset, plasma levels of GDF15 were measured in a longitudinal prospective study of children during T1D development (n=132 cases vs. n=40 controls) and in children with islet autoimmunity but normoglycemia (n=47 cases vs. n=40 controls) using targeted mass spectrometry. We also investigated the regulation of GDF15 production in islets by fluorescence microscopy and western blot analysis.

**Results:** The proximity-ligation assay identified ERBB2 as the GDF15 receptor in islets, which was confirmed using its specific antagonist, tucatinib. The untargeted proteomics analysis and caspase assay showed that ERBB2 activation by GDF15 reduces β cell apoptosis by downregulating caspase 8. In plasma, GDF15 levels were higher (p=0.0024) during T1D development compared to controls, but not in islet autoimmunity with normoglycemia. However, in the pancreatic islets GDF15 was depleted via sequestration of its mRNA into stress granules, resulting in translation halting.

**Conclusions/interpretation:** GDF15 protects against T1D via ERBB2-mediated decrease of caspase 8 expression in pancreatic islets. Circulating levels of GDF15 increases pre-T1D onset, which is insufficient to promote protection due to its localized depletion in the islets. These findings open opportunities for targeting GDF15 downstream signaling for pancreatic β cell protection in T1D and help to explain the lack of natural protection by the endogenous protein.

## 1. Introduction

Diabetes is a severe chronic metabolic disease that affects approximately 537 million people worldwide, being about 10% and 85% of the cases type 1 diabetes (T1D) and type 2 diabetes (T2D), respectively [1]. T1D is caused by an autoimmune destruction of the pancreatic β cells, while T2D is characterized by insufficient insulin secretion and insulin resistance in peripheral tissues [2]. Despite mechanistic differences, β-cell stress and insulin insufficiency are common underlying factors [3], open the possibility of developing therapies to target both diseases. In this context, growth/differentiation factor 15 (GDF15, also known as MIC-1, placental TGF-β, NAG-1, and NRG-1) has gained visibility as a potential common therapeutic target for both T1D and T2D [4].

GDF15 controls whole-body metabolism by regulating food intake, insulin sensitivity, and body weight [5]. Mechanistically, GDF15 activates the GFRAL receptor (GDNF family receptor ɑ-like) in the hindbrain, which controls appetite [6]. Circulating GDF15 levels are elevated in individuals with T2D [7]. GDF15 improves insulin secretion [8] and is involved in the mechanism of action of metformin [9], one of the most prescribed drugs for treating T2D. Therefore, GDF15 has been investigated as a therapeutic target for treating metabolic diseases and obesity. However, the mechanisms by which GDF15 improves insulin secretion in the pancreas are only partially known, as the GFRAL receptor is not normally expressed in pancreas [10, 11]. Additionally, GDF15 administration can cause nausea and anorexia as side effects [12], which is a significant challenge for developing it into a therapy.

We have reported that GDF15 production in human islets is reduced by pro-inflammatory cytokines and is decreased in islets from non-obese diabetic (NOD) mice with insulitis and individuals with T1D [13]. We also found that GDF15 is an islet protective factor, reducing cytokine-induced apoptosis, preventing insulitis, and reducing T1D onset in NOD mice [13]. How GDF15 exerts its protective effects remains elusive. A deeper understanding of its signaling network may identify therapeutic targets. Here, we address the mechanisms underlying GDF15 action (1) using molecular and pharmacological assays to identify a GDF15 receptor in islets; and (2) conducting proteomic analysis to determine the downstream protective signaling triggered by GDF15. We also studied why the endogenously produced GDF15 fails to provide protection against T1D development by measuring its levels in circulation and locally in islets. We performed targeted proteomics to quantify the levels of GDF15 in plasma prior to the onset of T1D and microscopy and immunological experiments to study its production in islets.

## 2. Methods

### 2.1. Tissue and cell culture

Human islets were received from the Integrated Islet Distribution Program (IIDP) or the Alberta Islet Isolation Center (details in **Table S1**), recovered overnight and hand-picked for experiments [13]. EndoC-βH1 human cell line was a kind gift from Dr. R. Scharfmann (Institut Cochin, University of Paris, France) [14], and cultured in Matrigel-fibronectin-coated plates [13]. Cultures were pre-treated with or without recombinant GDF15 (rGDF15, 100 ng/mL; ProSpec); for 24 h and treated with human IL-1β (50 U/mL) + human IFNγ (1000 U/mL) (R&D) for 24 h. Cultures were washed twice with 1X PBS and stored at -80°C for the downstream analysis. Cultures were screened regularly for mycoplasma contamination using the PCR Mycoplasma Detection Kit (ABM).

### 2.2. Proximity Ligation Assay (PLA)

PLA was performed on human pancreatic tissue obtained from nPOD biorepository with Duolink *In Situ* Red Starter kit Mouse/Rabbit (Sigma), according to the manufacturer’s protocol. After dehydration and antigen retrieval, slides were blocked with Duolink blocking solution at 37 °C for 1 h and incubated with the following primary antibodies: Rabbit anti-GDF15 (Bioss), mouse anti-ERBB2 (Thermo Scientific), and guinea pig anti-insulin (Dako). Images were acquired using a Zeiss LSM 800 confocal microscopy attached to an Airyscan detector (Zeiss, Germany). PLA images were processed by spot finding algorithm (Laplacian of Gaussian) and cytoplasmic signals were considered as interaction events between GDF15 and ERBB2. Nuclei were identified with manual annotation, and their masks were extended for estimating the cytoplasmic area. β cells were identified using insulin as a marker.

### 2.3. Western blot analysis

Islets were lysed with 50 mM of Tris-HCl, 150 mM NaCl, 0.05% deoxycholate, 0.1% IGEPAL, 0.1% SDS, 0.2% sarcosyl, 5% glycerol, 1 mM DTT, 1 mM EDTA, 2 mM MgCl_2,_ protease inhibitor (Complete mini, EDTA free, Roche) and phosphatase inhibitor (PhosphoStop, Roche), resolved by SDS-PAGE under denaturing conditions, and transferred onto a PVDF membrane. Membranes were blocked with Intercept Blocking Buffer (LI-COR) and incubated overnight at 4°C with the following primary antibodies: anti-Jak2 (Invitrogen), anti-phospho-Jak2, anti-STAT2 (Santa Cruz Biotechnology), anti-phospho-STAT2 (Cell Signaling), anti-caspase 8 (Proteintech), and anti-β-actin (Millipore). Membranes were incubated with respective secondary antibodies for 2 h. Membranes were imaged using an ODESSY CLx (LI-COR) and protein was quantified using Image Studio software (LI-COR).

### 2.4. Apoptosis assay

EndoC-βH1 cells were treated with or without 50 μM Z-IETD-FMK (Selleckchem) or 8 nM tucatinib (Selleckchem) in combination with rGDF15 (100 ng/mL), IL-1β and IFNγ for 48 h. Apoptosis was measured using a caspase-Glo 3/7 assay (Promega), following manufacturer recommendations, for 3 h with 30 min intervals. The time point with the highest signal intensity was chosen to calculate the caspase 3/7 activity. Statistical analysis was done with GraphPad Prism10.

### 2.5. Untargeted proteomics of human islets

Islets were digested with trypsin and labeled with a 16-plex TMT kit (Thermo Fisher Scientific) as previously described [13]. Peptides were multiplexed, fractionated by high pH reverse phase chromatography, and analyzed by LC-MS/MS on an Acquity M-Class Nano UHPLC system (Waters) connected to a Q-Exactive mass spectrometer (Thermo Fisher Scientific) [13]. Data were processed with MaxQuant [15], by searching against a human SwissProt database (April 12, 2017, 20,198 sequences). The default settings for precursor and fragment mass tolerance were used. Peptide searching was performed with specific trypsin digestion with up to two missed cleavage sites. Carbamidomethylation of cysteine was set as a fixed modification; acetylation of protein N-terminus and oxidation of methionine residues were set as variable modifications. The false discovery rate (FDR) was set to 1% at peptide and protein levels. Data were log2 transformed, controlled for quality, normalized to total abundance, and submitted to statistical analysis with a standard Analysis of Variance (ANOVA) model [16]. Network analysis was performed with Ingenuity Pathway Analysis software (Qiagen). GraphPad Prism 10 and Cytoscape v3.9.1 were used for graphical representation. Heatmaps were generated using the k-mean clustering function in Perseus-MaxQuant.

### 2.6. Targeted proteomics of GDF15 in human plasma

GDF15 measurement was conducted on samples from the Diabetes Autoimmunity Study in the Young (DAISY) study in parallel with the analysis of complement proteins [17]. DAISY has monitored 2,547 children at increased risk for T1D, consisting of first-degree relatives of individuals with T1D and children with T1D susceptibility HLA *DR-DQ* genotypes [18]. We analyzed plasma samples from 172 children (40 control and 132 cases with 48% and 40% female, respectively) collected over time up to the age 23 years, including samples from 47 and 131 children collected prior and after the appearance of autoantibodies (seroconversion), respectively. Protocols approved by the Colorado Multiple Institutional Review Board and written informed consent was obtained from donors or their parents. Proteins were digested with trypsin in an Eppendorf epMotion 5075 Liquid Handler, and peptides were analyzed on an Acquity M-Class Nano UHPLC system (Waters) connected to a triple quadrupole mass spectrometry (TSQ Altis, Thermo Fisher) as previously described [17]. Data were extracted with Skyline software [19] with peak boundaries and alignment manually inspected, and submitted to quality control [16], followed by statistical analysis using a linear mixed model comparing the control group to cases prior to or after seroconversion. The analysis was adjusted for sex, HLA group, and first-degree relative status with a nested random effect for subject and plate. For visualization purposes, the data was averaged to peptide abundance within group for five age ranges: (1) <3 years, (2) 3 to 6 years, (3) 6-9 years, (4) 9-12 years, and (5) >12 years.

### 2.7. Fluctuation localization imaging-based fluorescence in situ hybridization (fliFISH) and direct Stochastic Optical Reconstruction Microscopy Immunofluorescence (dSTORM IF) staining

EndoC-βH1 cells were seeded in a chamber slide (75,000 cells/well) (Lab-Tek II and were treated as described above. Slides were fixed with 4% PFA for 10 minutes at room temperature, washed 3x with PBS, and super-resolution imaging for mRNA and protein targets was performed with fliFISH and dSTORM, respectively [20]. Images were collected with a single-molecule microscope (Olympus IX71 equipped with 405 nm, 488 nm, 542 nm, and 640 nm solid lasers) with 100× oil immersion objective lens (NA 1.4) and an EMCCD camera (Andor iXon Ultra 897). Data processing was performed with in-house developed MATLAB and C scripts. For fliFISH, the Alexa647-labeled secondary single-probe on-time fraction was characterized to be 0.4%. Assuming a 70% hybridization efficiency, a 4% ensemble on-time fraction was set as the threshold for GDF15 transcript detection, which was centered to the mass center. dSTORM images were collected with 20 Hz frame rate, 1 kW/cm^2^ laser power and >10,000 frames. Photoswitching was activated with GLOX-containing buffer: 50 mM Tris, 10 mM NaCl, 10% glucose, 560 µg/mL glucose oxidase, 34 µg/mL catalase and 1% β- mercaptoethanol. Cell boundaries were segmented based on their autofluorescence and DAPI staining. Detection threshold was set to 20-fold higher intensity than the fluorescence background, being a distance of “0” between transcripts and proteins considered co-localization.

### 2.8. Human Pancreas Analysis Program (HPAP) data processing

Single-cell transcriptomics raw FASTQ files were retrieved from HPAP PANC DB [21] and aligned against the hg38 refence using Cell Ranger (V6.1.1) [22]. “Ambient” mRNA and doublets were removed with SoupX (V1.6.1) [23] and scDbIFinder (v3.16), respectively, and data was filtered with nFeature_RNA >200 and <9000, percent of mitochondrial reads <15% and nCount_RNA <10000. Sequences were clustered using Seurat (v4.1.1) [24] against cell type marker genes (INS, GCG, SST, TTR, IAPP, PYY, KRT19 and TPH1). Cell types were annotated using scSorter (v0.0.2) [25] based on markers, dimensionality reduction and Uniform Manifold Approximation and Projecting (UMAP). Annotated β cells (10,167 cells) were extracted from the matrix for expression analysis and normalized with Seurat. The top 3000 variable genes were used for the Principal Component Analysis and integrated using Harmony (v0.1.1) [26].

## 3. Results

### 3.1. ERBB2 as a GDF15 receptor in islets

ERBB2 (HER2) is a GDF15 receptor in cervical cancer [27] and oral squamous carcinoma [28] cells. Importantly, ERBB2 is expressed in β cells [29] and GDF15 enhances insulin secretion in β cells in a GFRAL-independent manner [10]. We analyzed the expression of ERBB2 signaling components (its canonical ligand neuregulin 1 (*NRG1*), and the co-receptors *EGFR* (*ERBB1*), *ERBB3*, and *ERBB4*) in β cells in the single-cell HPAP transcriptomic dataset (**Table S2**). All components of the ERBB2 signaling complex are expressed at low levels in non-diabetic and autoantibody-positive individuals, but the expressions of *NRG1*, *ERBB1*, and *ERBB3* increased in β cells from individuals with T1D (**Figure 1A**). We performed a proximity ligation assay (PLA) on human pancreatic tissue sections to determine if GDF15 interacts with ERBB2 on the β cells (**Figure 1B, left panel**). A GDF15-ERBB2 interaction was observed, as indicated by the PLA signal foci detected in insulin-positive cells. Further quantification of the number of cytoplasmic PLA foci per cell indicates a significant increase in the foci number in diabetic compared to nondiabetic insulin-positive cells (**Figure 1B, right panel**).

**Figure 1.**
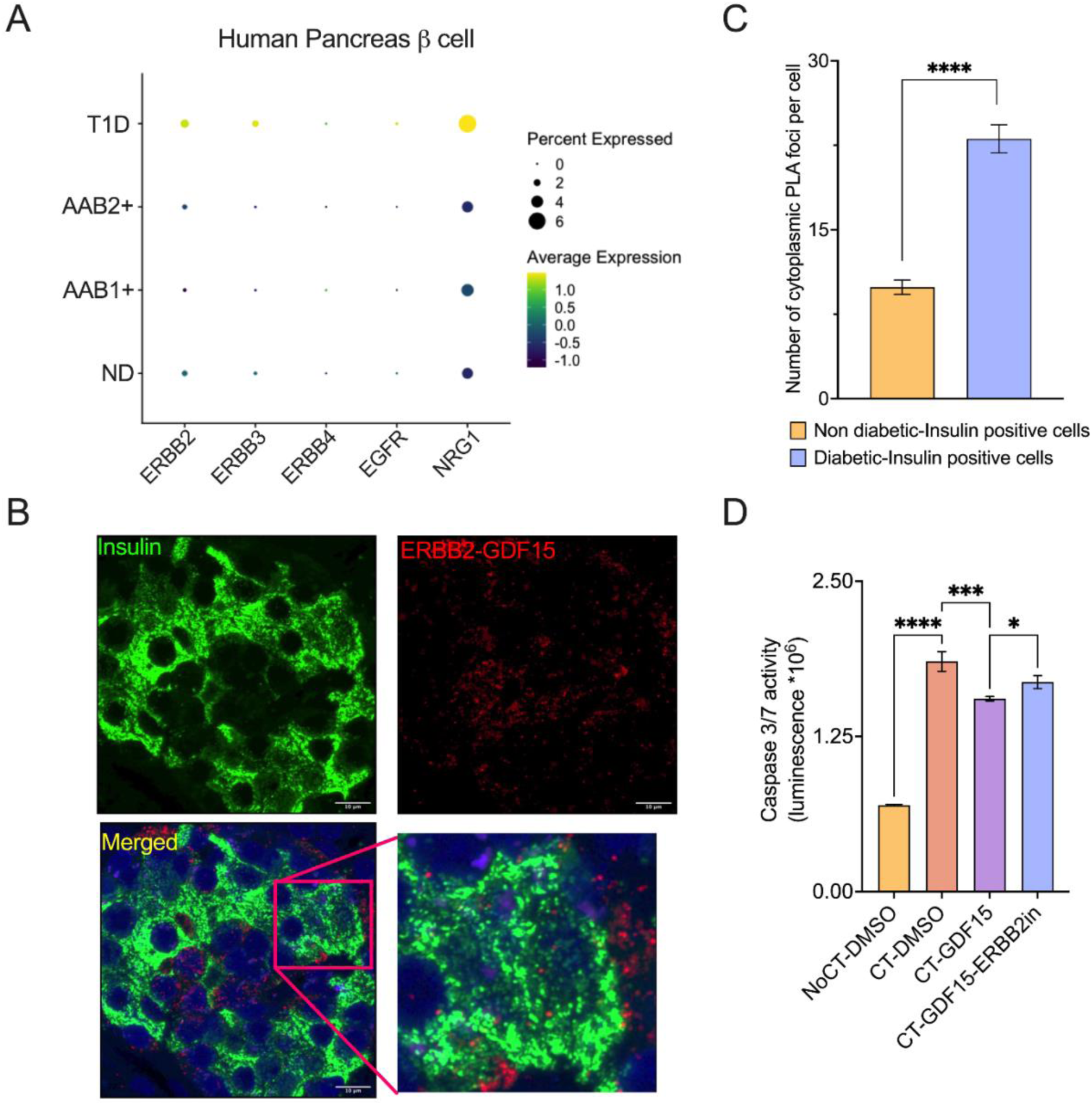
GDF15 binds to the ERBB2 receptor. **A.** Expression of ERBB1 (EGFR), ERBB2, ERBB3, ERBB4, and NRG1 in HPAP scRNA-seq dataset of human pancreatic islets. The size of the dots indicates the percentage of cells expressing the genes, while the color indicates the scaled average expression. **B.** Representative proximity ligation assay (PLA) data, showing ERBB2 and GDF15 foci (red) in insulin-positive islets cells (green). **C.** Quantitation of ERBB2-GDF15 foci in insulin-positive cells from nondiabetic (n=287) vs. diabetic (n=431) donors (Mean ± SD). **D.** Caspase 3/7 activity assay performed in EndoC-βH1 cells (n=3) treated with cytokines (IL-1β and IFNγ), GDF15, and ERBB2 inhibitor (tucatinib) (Mean ± SD). Mann Whitney test and one-way ANOVA were performed for C and D, respectively, with *p<0.05, ***p<0.001, and ****p<0.0001.

To investigate if ERBB2 signaling is required to protect β cells against cytokine-mediated apoptosis, we treated human EndoC-βH1 cells with recombinant GDF15 (rGDF15) in the presence or absence of the ERBB2 specific antagonist tucatinib and the cytokines IL-1β + IFN-γ for 48 h. Apoptosis was measured by the caspase 3/7 activity assay, which showed a decrease in cytokine-mediated activation of caspase 3/7 activity in the presence of rGDF15 (**Figure 1C**), confirming GDF15’s protective effect. Tucatinib reduced the protection against cytokine-induced apoptosis by rGDF15, indicating the role of ERBB2 in this signaling cascade. Overall, these results indicate that GDF15 regulates cytokine-mediated apoptosis by triggering ERBB2 signaling in β cells.

### 3.2. Identification of GDF15-regulated proteins in human islets

To elucidate the GDF15 signaling pathways, we performed untargeted proteomic analysis of human islets treated rGDF15 for 48 h (**Table S3**). rGDF15 treatment led to the regulation of 796 proteins compared to untreated control (**Figure 2A**), and ingenuity pathway analysis (IPA) showed enrichment of 145 biological pathways (**Table S4**). From those pathways, we mapped 108 pathways in which IPA provided a predicted activation or inhibition based on the enriched protein abundance (**Table S4**). We then consolidated these results into 91 main pathways that shared common proteins using the EnrichmentMap tool in Cytoscape (**Figure 2B**). The pathways with the most mapped proteins were the estrogen pathway, the cachexia signaling pathway, protein kinase A, hepatic fibrosis signaling pathway, and RAR activation, all having 22 mapped proteins. AMPK signaling and EIF2 signaling also were among the top hits of the list. Most of the inositol biosynthesis and degradation pathways were predicted to be strongly downregulated, whereas AKT-related pathways and ERK/MAPK signaling were predicted to be upregulated (**Figure 2B**). As GDF15 binds to the ERBB2 receptor, we mapped the GDF15-regulated proteins to the HER2 (ERBB2) signaling pathway using IPA. Both the PI3K/AKT and the MAPK pathway were predicted to be activated to either reduce apoptosis or increase cell survival/proliferation via the ERBB2/3 receptor (**Figure 2C**).

**Figure. 2.**
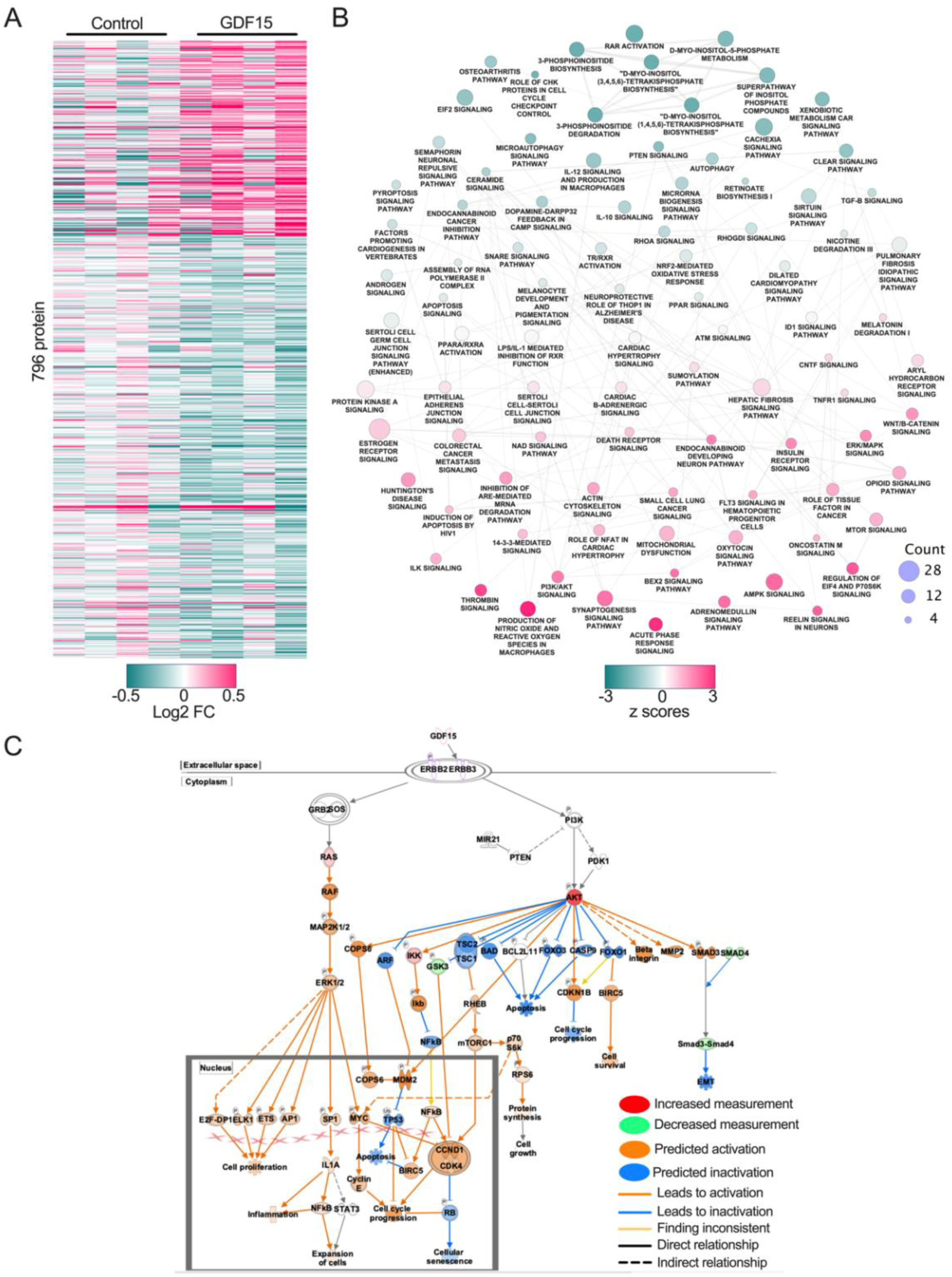
Downstream GDF15 signaling. **A.** Heatmap showing 796 differentially regulated proteins by GDF15. Data represented as Log2 fold-change (FC) compared to control. **B.** Nodes and edges plot showing the significantly enriched pathways (91 pathways) determined using ingenuity pathway analysis (IPA) software. The size of the node represents the number of proteins enriched, while the color (denoted as z scores) represents the predicted activation and inactivation of the respective pathways. The thickness of the edges represents the similarity coefficient (0-6) calculated by the EnrichmentMap tool in Cytoscape. **C.** Enrichment of GDF15-regulated proteins onto the ERBB2 (HER2 signaling in breast cancer) signaling pathway using IPA. The unenriched section of the pathway map was trimmed.

To contextualize the effect of GDF15 under inflammatory conditions, we analyzed the differential proteomic changes in IL-1β + IFN-γ-treated human islets with or without rGDF15. We identified 532 proteins that were significantly modulated by GDF15 in this condition (**Figure 3A**). We mapped these proteins to biological functions using IPA and identified 25 statistically significant pathways. Cell-death and cell-survival pathways were the most enriched with 214 proteins, being 148 proteins mapped to cell survival, 112 proteins to apoptosis functions with 74 proteins shared between these functions (**Figure 3B-C, Table S5**). Our analysis showed that the cell survival was predicted to be activated while the apoptosis inactivated (**Figure 3C**). Together, these findings indicate that GDF15 plays antiapoptotic and cell survival roles in islets in both normal and inflammatory conditions.

**Figure 3.**
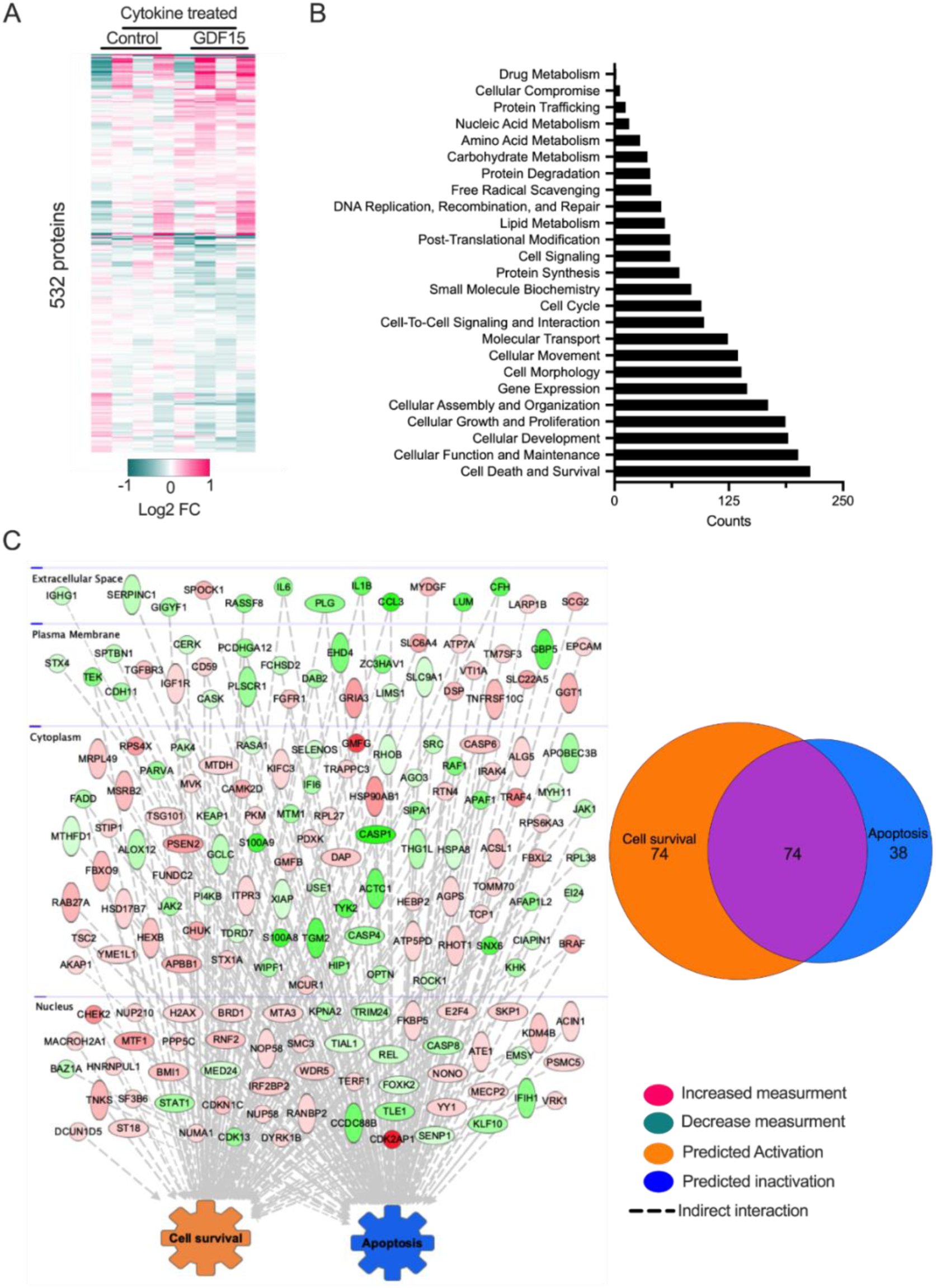
GDF15-mediated functions in the presence of cytokines. **A.** Heatmap representing GDF15-mediated differentially expressed proteins (532 proteins) in islets in the presence of cytokines (IL-1β + IFNγ). Data represented as Log_2_ FC are compared to the cytokine-treated control group. Islets were pre-treated with GDF15 for 24 h prior to 24 h treatment with cytokines. **B.** Bar graph representing the molecular and cellular functions enriched by differentially expressed proteins from panel A. **C.** Left panel: all the proteins, with their respective FC measurement, mapped to cell survival and apoptosis molecular functions. Right panel: Venn diagram showing the number of proteins that are shared between the two functions.

### 3.3. GDF15 regulates caspase 8 to reduce cytokine-mediated apoptosis

We analyzed the islet proteomics data to identify proteins of the ERBB2-GDF15 signaling that are responsible for anti-apoptotic effects. We identified 76 proteins that were modulated by both cytokine and GDF15 treatments, being 37 of these proteins mapping to the cell death and survival biological functions (**Figure 4A**). The pathway analysis determined that these proteins participate in the inactivation of apoptosis through JAK2, STAT1, and caspase 8 proteins that are directly regulated by the GDF15-ERBB2 signaling cascade via PI3K/AKT and ERK/MAPK pathways (**Figure 4B**). Western blot analysis of human islets confirmed that both the phosphorylated states of STAT2 and JAK2 and the protein expression levels of STAT2, JAK2, and caspase 8 are modified by cytokines. Moreover, GDF15 treatment impacted the expression of these proteins, particularly causing a reduction in the cytokine-induced expression of caspase 8 (**Figure 4C**). In agreement with these findings, there was an increase in *CASP8* and *JAK2* gene expression in human β cells from individuals with T1D compared to control donors (**Figure 4D**). To determine the role of caspase 8 in GDF15-mediated antiapoptotic activity, we treated EndoC-βH1 cells with cytokines and a caspase 8 inhibitor (Z-IETD-FMK) for 48 h. Caspase 8 inhibition significantly blocked cytokine-mediated apoptosis (**Figure 4E**), indicating that GDF15 mediates its protective effect by reducing the abundance of caspase 8 protein in islets.

**Fig. 4.**
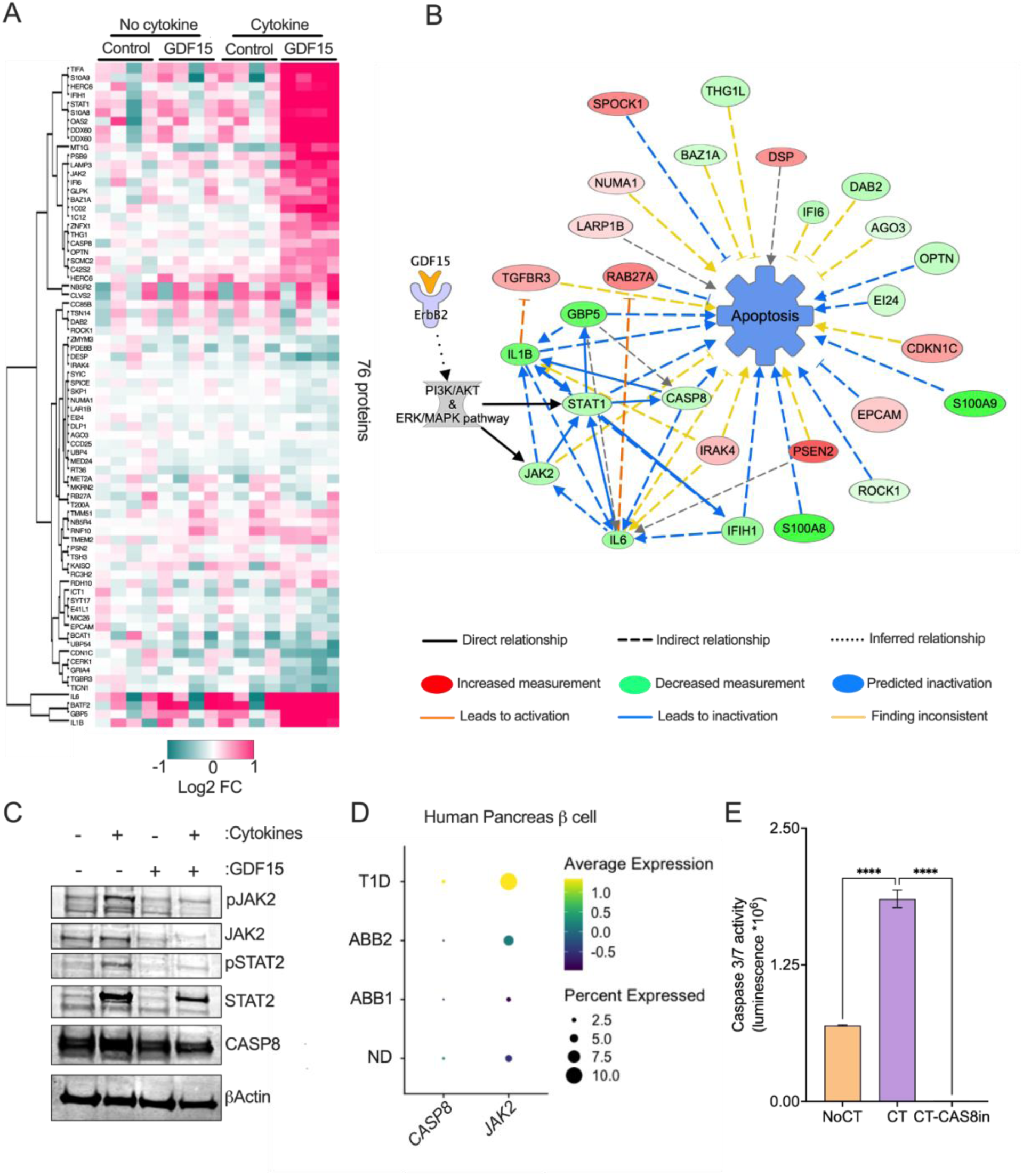
GDF15 regulates caspase 8-mediated β cell protection. **A.** Heatmap showing 76 proteins that are significantly affected by cytokines as well as by GDF15. Data represented as log2 fold-change compared to untreated control. Islets were pre-treated with GDF15 for 24 h prior to 24h treatment with cytokines. **B.** Node and edge plot generated by IPA that represents all the proteins (from A, right panel) that are associated with the inactivation of the apoptosis functions in human islets by rGDF15 treatment. **C.** Western blot image showing protein abundance of pJAK2, JAK2, pSTAT2, STAT2, and CASP8 in islets treated with cytokines and GDF15 for 24 h. **D.** CASP8 and JAK2 expression from scRNA-seq analysis of human pancreatic islets from the HPAP dataset. The size of the dots indicates the percentage of cells expressing the genes, while the color indicates the scaled average expression. **E.** Caspase 3/7 activity assay performed in EndoC-βH1 cells (n=3) treated with cytokine (IL1β + IFNγ) and CASP8 inhibitor (CASP8in: Z-IETD-FMK) for 24 h. One-way ANOVA was performed for E, with *p<0.05 and ****p<0.0001.

### 3.4. GDF15 levels are elevated prior to T1D development

We measured the circulating levels of GDF15 prior to T1D development using targeted proteomic analysis in plasma of the longitudinal, prospective study DAISY. Samples from individuals who developed T1D during the study (n=132 cases vs. n=40 controls), individuals who developed islet autoimmunity but remained normoglycemic (n=47 cases vs. n=40 controls), and age-matched healthy controls were used (**Table S6**). We found that two GDF15 peptides were significantly increased (p=0.0024 and p=0.04) in plasma from individuals who developed T1D compared to matched controls (**Figure 5A**). However, the levels of GDF15 were not affected in normoglycemic-islet autoimmunity positive individuals (**Figure 5B**). These results show an increase in GDF15 prior to the T1D onset.

**Figure 5.**
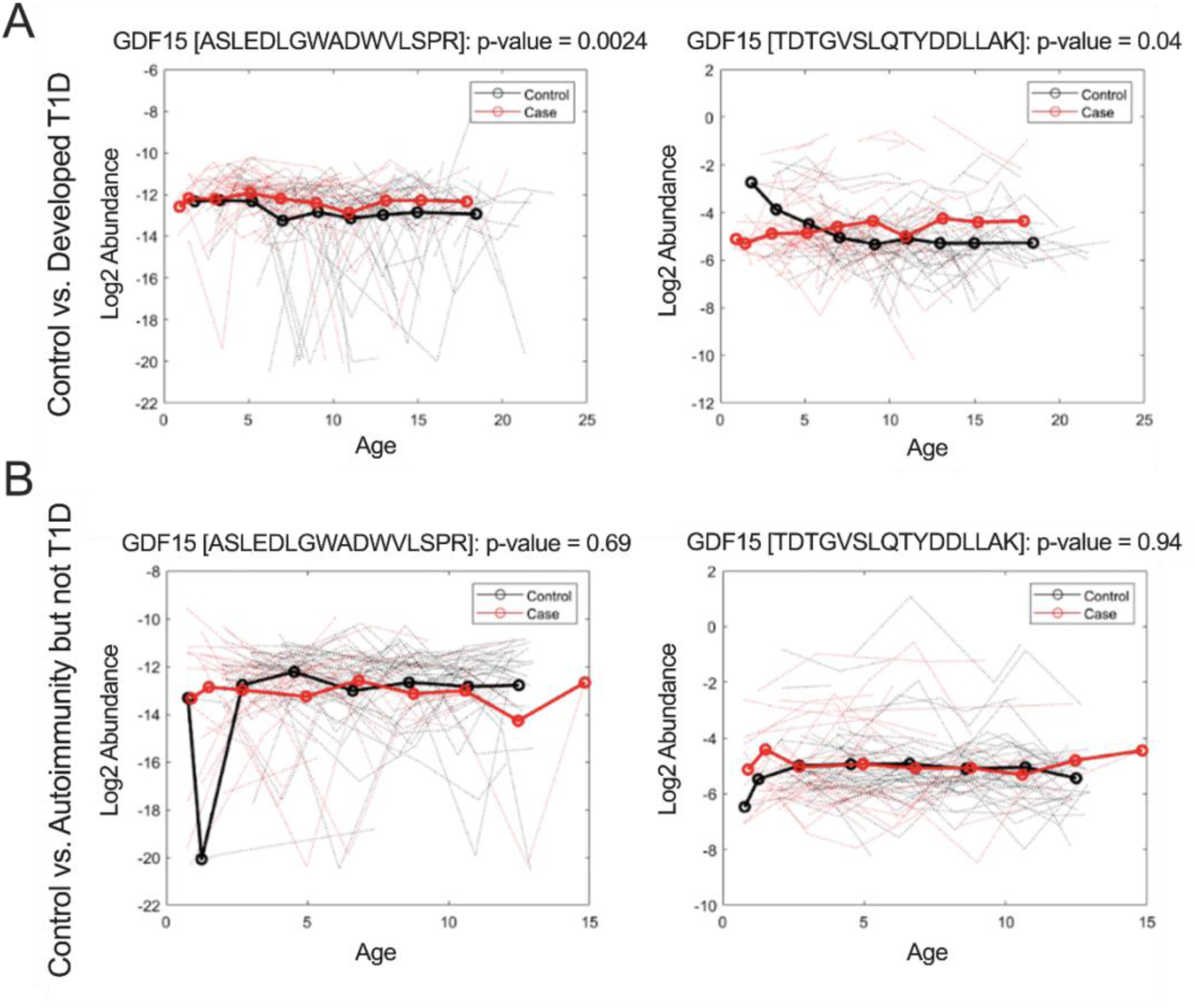
Targeted mass spectrometry analysis in Diabetes Autoimmunity Study in the Young (DAISY) cohort shows an increased level of two GDF15 peptides in the group that developed T1D. **A.** & **B.** Shows log2 abundance of two GDF15 peptides (ASLEDLGWADWVLSPR and TDTGVSLQTYDDLLAK) over time in children with autoimmunity that developed to T1D (n=131 vs. n=40 controls) or stayed normoglycemia (n=47 vs. n=40 controls) for the period of the study, respectively. P value was calculated using a linear mixed effects model.

### 3.5. Targeting of GDF15 pre-mRNA to stress granules in insulitis

We investigated the expression and mechanism of GDF15 regulation in pancreatic islets. We previously reported that GDF15 mRNA translation is blocked in EndoC-βH1 cells and human islets treated with IFNγ + IL-1β [13]. Here, we investigate the mechanism of this translational blockage. GDF15 mRNA contains an AU-rich element (ARE), which is a binding site for the RNA-binding protein tristetraprolin (TTP, **Figure 6A**). TTP targets mRNAs to stress granules [30] and cytokines can induce stress granule formation [31]. Upon cytokine treatment TTP transcript, protein, and phosphorylation were upregulated (**Figure 6B-C**). We next monitored GDF15 mRNA association with stress granules in EndoC-βH1 cells treated with IL-1β + IFN-γ for 24 h by visualizing GDF15 transcript, TTP protein, and the stress granule marker G3BP1 using super-resolution STORM microscopy (**Figure 6D-E**). There was an increased association between GDF15 mRNA with G3BP1 and TTP in cells treated with cytokines, indicating that translationally inhibited GDF15 mRNAs are diverted to stress granules during β cell stress.

**Figure 6.**
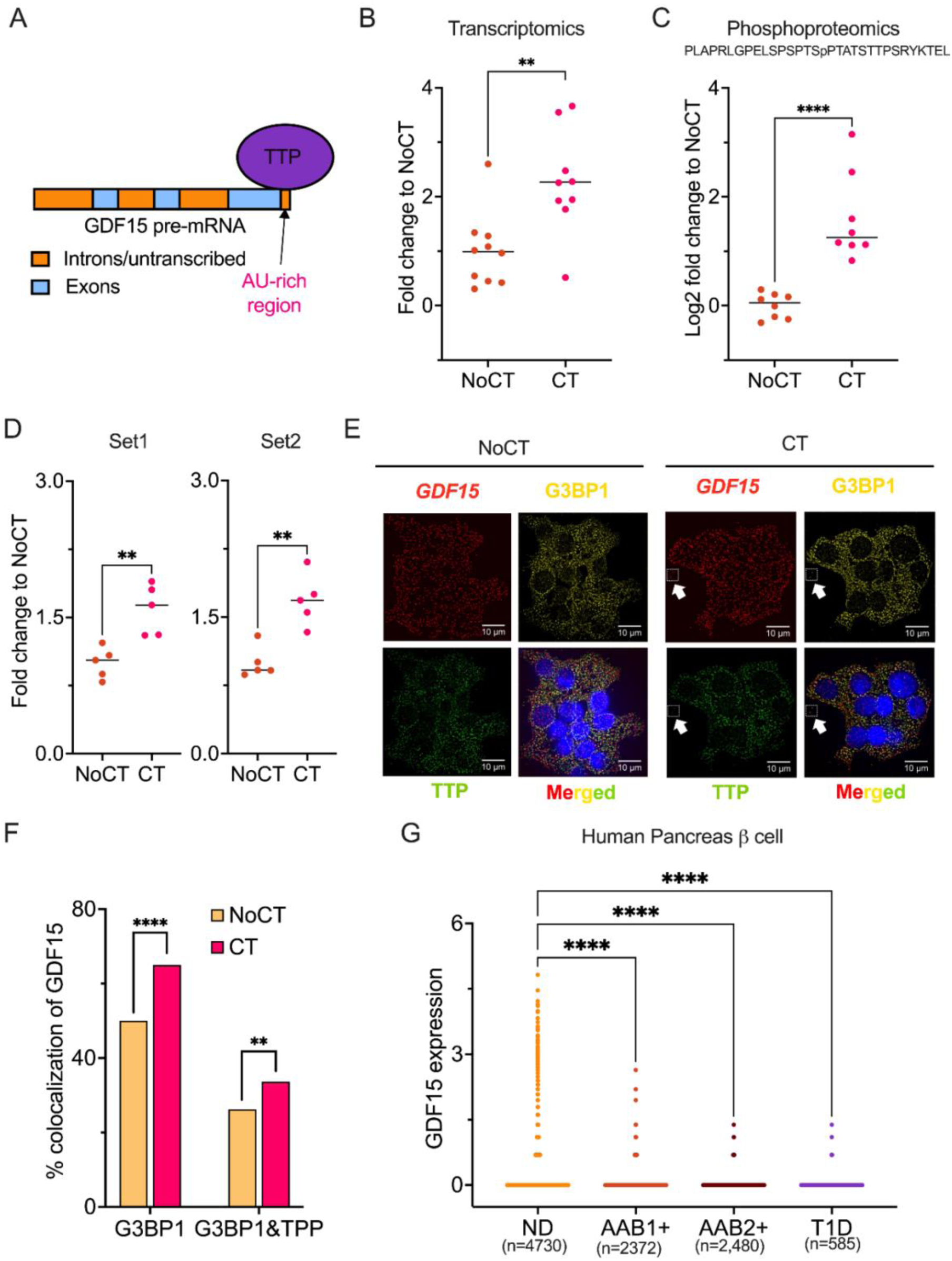
Targeting of GDF15 pre-mRNA to stress granules in β cells. **A.** Schematic diagram showing TTP-mediated regulation of GDF15 via binding to the AU-rich pre-mRNA region in GDF15. **B. & C.** Reanalyzed TTP transcriptomics (n=10) and phosphoproteomics (n=8) levels in human islets treated with or without cytokine (IFN-γ and IL-1β treatment for either 48 h or 24 h) from Ramos-Rodriguez et al. [49] and Yi et al. [50], respectively. The measured phosphopeptide sequence is listed in the figure with the modified site represented by “p”. **D.** TTP protein level from reanalyzed proteomics data from Nakayasu et al. [13], in human islets (set 1 and 2, n=5 each) treated with or without cytokine (24 h, IFN-γ and IL-1β treatment). **E.** Representative image showing the co-localization of *GDF15* mRNA (FISH), G3BP1 (IF), and TTP (IF) in EndoC-βH1 cells treated with/without cytokines (IL-1β and IFNγ) for 24 h. **F.** Quantitation of E., represented as % colocalization of *GDF15* with G3BP1 or G3BP1 and TPP, in presence and absence of cytokine. **G.** scRNA-seq dataset of human pancreatic islets, downloaded from Human Pancreas Analysis Program (HPAP) (https://hpap.pmacs.upenn.edu). N=9 for T1D, n=8 for AAB1+ (islet autoimmunity, each donor is positive for one autoantibody), n=2 for AAB2+ (islet autoimmunity, each donor has >=2 autoantibodies), and n=16 for non-diabetic (ND) individuals. Student’s t-test and one-way ANOVA were used to calculate significance in C & D, respectively, where **p<0.01 and ****p<0.0001.

TTP targets transcripts to stress granules but in the long term it induces the degradation of ARE-containing mRNAs [30]. We monitored GDF15 mRNA expression in β cells during T1D development in the single-cell transcriptomics data [21] (**Figure 6F**). Expression of the *GDF15* transcript was highest in β cells from control donors, with a gradual reduction in individuals with islet autoimmunity (characterized by the presence of 1 (AAB1+) or 2 (AAB2+) autoantibodies against the islets) and established T1D. These results show that despite GDF15’s increase in circulation during T1D development, the local islet levels are drastically reduced, probably due to the targeting of GDF15 mRNA to stress granules by TTP.

## 4. Discussion

### 4.1. Defining GDF15 receptor and downstream signaling in β cells

GFRAL is the GDF15 receptor in the hindbrain, working in combination with the coreceptor Rearranged during Transfection (RET) [6]. In pancreas, this receptor expression is normally at trace levels and although it is increased in pancreatic ductal carcinoma [11], GFRAL is unlikely to be a receptor for GDF15 in this tissue. GFRAL is not responsible for GDF15-mediated insulin secretion [10]. ERBB2, CD44 and CD48 have been recently described as GDF15 receptors [27, 28, 32, 33], although their role in islets remains to be investigated. Here, we identified ERBB2 as a receptor for GDF15 in β cells. ERBB2 is a receptor tyrosine kinase family member with structural similarity with the epidermal growth factor receptor, and heterodimerize with ERBB1 (HER1 or EGFR), ERBB3 or ERBB4. Genome-wide association studies (GWAS) have identified ERBB3 polymorphism as a T1D risk factor [34]. GDF15 binds to ERBB2 causing breast-cancer resistance to Herceptin, a drug that blocks the activation of this receptor by neuregulins [35, 36]. Moreover, there is a negative correlation between breast cancer and T1D, but the role of ERBB2 and GDF15 in this relationship has not been investigated [37].

In the downstream signaling, we found that GDF15 downregulates metallothionein-1G, a member of the small cysteine-rich proteins that play a role in metal homeostasis. Metallothionein-1 negatively regulates the glucose-stimulated insulin secretion [38] and its reduction by GDF15 may improve insulin secretion. The downstream GDF15-ERBB2 signaling also suppresses the expression of caspase 8, preventing β-cell apoptosis. Similarly, in cancer cells, ERBB2 inhibits apoptosis by downregulating the expression of caspase 8 [39]. Caspase 8 plays an important role in amyloid-induced β cell apoptosis in both humans and mice [40] and - in line with our findings - GDF15 promotes optic neuron survival following crush by suppressing the expression of caspase 8 [41].

The identification of the mechanisms by which GDF15 protects β cells against pro-inflammatory cytokines opens opportunities for targeting the GDF15 pathway for therapeutic development. Metallothionein is a therapeutic target for inflammatory bowel disease [42], as well as neuregulin and ERBB2 mimetics for various cardiac issues [43, 44]. These drug candidates can be repurposed and should be tested for T1D too. Caspase 8 is a drug target for cancer [45] but in the context of increasing its expression, which is the opposite wanted function for T1D pathology. Further, there is a significant lack of research on ERBB agonist space, and our study warrants a second look at finding a novel therapy targeting ERBB2 and caspase 8 to mimic GDF15 effect in β cells.

### 4.2. Local vs. circulating levels of GDF15

GDF15 levels are increased in multiple conditions, including T2D, high-intensity exercise, and cardiac arrest [46]. Similarly, we found an increase in the level of GDF15 prior to T1D onset. There is also an increase of GDF15 expression in β cells under endoplasmic reticulum stress conditions [47]. However, the level of GDF15 in pancreatic islets is reduced in NOD mice with insulitis and depleted in humans with T1D [13]. GDF15 mRNA levels increase while the protein levels decrease post-cytokine treatment in human islets, indicating a post-transcriptional regulation [13]. Here, we provide an explanation for this paradox by showing that despite GDF15 mRNA is induced by cytokines, they are sequestered in the stress granule, blocking its translation. Further, the single-cell transcriptomics data show a depletion of GDF15 mRNA in chronic T1D conditions, which might be a subsequent step of GDF15 regulation. As TTP plays different roles in mRNA expression, localization, and degradation depending on its phosphorylation state by different protein kinases [48], it is possible that TTP blocks GDF15 production not only targeting its mRNAs to stress granules, but also by further targeting it to degradation. Overall, despite the increase in the circulating level of GDF15, a reduction in local levels prevents its beneficial effects against T1D development.

### 4.3. Conclusions

We found that GDF15 engages the ERBB2 receptor in islets instead of GFRAL receptor as described in the hindbrain. Because a major side effect of GFRAL activation by GDF15 is the induction of nausea, this finding opens perspectives for specifically targeting GDF15 or its downstream signaling in β cells to develop anti-diabetic drugs. ERBB2 is a main target of therapies for breast cancer, and a variety of tools are available for drug screening and development, which could accelerate the development of anti-diabetic drugs targeting ERBB2. We also showed that circulating GDF15 level is increased during the development of T1D. However, these increased systemic levels are insufficient to prevent β cell loss due to depletion of GDF15 locally in the pancreatic islets. Therefore, a targeted drug that restores GDF15 levels in β cells also hold potential as therapeutic target.

## Acknowledgments

The authors thank Jacob. R. Enriquez for his help with data analysis. The authors also thank the NIDDK-supported Integrated Islet Distribution Program (IIDP) for providing the human islets used in the study. Part of the work was performed in the Environmental Molecular Sciences Laboratory, a U.S. Department of Energy (DOE) national scientific user facility at Pacific Northwest National Laboratory (PNNL) in Richland, WA. Battelle operates PNNL for the DOE under contract DE-AC05-76RLO01830. This research was performed with the support of the Network for Pancreatic Organ donors with Diabetes (nPOD; RRID:SCR_014641), a collaborative type 1 diabetes research project supported by JDRF (nPOD: 5-SRA-2018-557-Q-R), and The Leona M. & Harry B. Helmsley Charitable Trust (Grant#2018PG-T1D053, G-2108-04793). The content and views expressed are the responsibility of the authors and do not necessarily reflect the official view of nPOD. Organ Procurement Organizations (OPO) partnered with nPOD to provide research resources that are listed at http://www.jdrfnpod.org/for-partners/npod-partners.

## Data Availability

The raw mass spectrometry data can be found at https://massive.ucsd.edu (targeted: MSV000090848, untargeted: MSV000093466) and the processed targeted mass spectrometry data is in Skyline (https://panoramaweb.org/DAISY_SRM_PNL.url).

## Supplementary files

Supplementary tables, files and checklists were uploaded to Open Science Frame. DOI 10.17605/OSF.IO/JDGNR

